# Assessing the Impact of Interventions on Tuberculosis Control: India Based Modelling Framework

**DOI:** 10.64898/2026.05.20.26353466

**Authors:** Yashi A. Raj, Raghavan Parthasarathy, Mithun Mitra, Sarika Mehra

## Abstract

**Background:** India accounts for nearly one-fourth of the global tuberculosis (TB) burden. The country’s progress towards elimination of TB is hindered by considerable heterogeneity in behavioural, social, and health system determinants, which influence transmission dynamics and care access. Evidence from the recent national TB prevalence survey showed that almost half of individuals with active disease were asymptomatic, underscoring the limitations of symptom-based case finding. Achieving the End TB targets will therefore require strategies that simultaneously address the substantial pool of individuals with undiagnosed, asymptomatic disease and those symptomatic individuals who do not seek care.

**Methods:** We developed a transmission model of TB that explicitly incorporates individuals with asymptomatic disease, and those who do not seek care. Model calibration was performed within a Bayesian framework using epidemiological and programmatic data for India. The calibrated model was then used to project the potential impact of intervention on TB incidence and mortality.

**Results:** Under the baseline scenario, the estimated TB incidence and mortality rates for 2024 were 180 (163-203) and 24 (18–31) per 100,000 population, respectively. Across all intervention scenarios targeting improved diagnosis, active case finding, nutrition support and their combination the reduction in incidence rate by 2030 ranged from 13% to 60% compared with 2025, while the corresponding decline in mortality rate ranged from 16% to 66%.

**Conclusion:** While individual interventions yield measurable reductions in TB incidence and mortality, but greater impact is achieved when implemented in combination reflecting the need for a comprehensive, multi-component response towards TB elimination.

**What is already known on this topic:**

- Studies have analysed the impact of single intervention scenarios across different regions and India

**What this study adds:**

- We developed a dynamic model of tuberculosis in India, modelling the impact of four different interventions: nutritional support, health facility improvements, active case finding and a combination of all three.
- We found that a single intervention cannot be considered powerful enough to improve the rate of decline of incidence and mortality and so a multi-sectoral intervention scenario is required.

**How this study might affect research, practice or policy:** Accelerating the decline in tuberculosis burden requires interventions targeting the entire care cascade while simultaneously providing nutritional support.

## INTRODUCTION

India has made substantial progress in reducing the burden of tuberculosis (TB) over the past decade. Between 2015 and 2024, TB incidence declined by 21%, compared to a global decline of 12%, while TB mortality decreased by 28% during the same period (1). Despite these gains, India continues to carry the largest global TB burden, accounting for approximately 27% of the world’s TB cases. In absolute terms it translates to over 2.7 million incident cases and more than 300,000 TB related deaths in 2024 (1). Achieving substantial reductions in the coming years requires accelerating the decline in incidence and mortality in comparison to the current trends and generating evidence on the relative impact of scale up of targeted interventions to meet India’s END-TB targets of reducing incidence by 80% and mortality by 90% compared to 2015.

TB, caused by Mycobacterium tuberculosis (Mtb), has been considered to follow the natural history of progression to disease through a latent, non-infectious phase of infection and an active, symptomatic, and infectious phase of disease. However, recent findings (2,3) have challenged this paradigm of active disease, identifying individuals with TB disease go through a stage where they remain asymptomatic. This group of individuals with asymptomatic TB disease were estimated to constitute a higher proportion of those remaining undetected across the world (4), prompting its recognition as an intermediate subclinical or asymptomatic category within the active TB disease paradigm (5,6).

With a network of public and private healthcare providers distributed across the country, pathways to TB diagnosis are complex and often prolonged. Individuals affected with TB disease and having signs and symptoms of TB frequently move between informal providers such as chemists and traditional healers and formal providers including hospitals and clinics before receiving a definitive diagnosis (7,8). This care-seeking pattern, combined with strained diagnostic infrastructure, contributes to substantial delays in treatment initiation and undermines TB control. The challenge is further compounded by individuals with asymptomatic TB, who are less likely to seek care and therefore remain undiagnosed for extended periods (9). These individuals continue to engage in routine community interactions, acting as potential reservoirs of infection, while also escaping detection by symptom-based screening strategies. In this study, we aim to generate evidence by quantifying the contribution of asymptomatic TB to transmission dynamics and estimating the overall disease burden through a transmission modelling framework.

## METHODS

We developed a compartmental model of TB transmission that incorporates the natural history of the disease, the care-cascade and the post treatment outcomes of relapse and reinfection. It has been assumed that individuals affected with TB disease develop symptoms after remaining asymptomatic for a period of time. The model also accounts for individuals who remain outside the formal care-seeking pathway (i.e. public and private providers engaged with the National TB Elimination Programme). Such a component in the modelling framework allows us to capture the epidemiological role of individuals who delay or forego care, and to more realistically represent the care cascade pathways which people adopt.

### Model description

The model represents TB natural history through five core states: uninfected *U*, two latent infection states (fast *L*_*f*_ and slow *L*_*s*_), and two TB disease states (asymptomatic *S*_*c*_ and symptomatic *A*). Following infection, individuals enter the latent fast compartment, from which they may stabilize into a latent slow state or progress to asymptomatic TB (*S*_*c*_). Reactivation of individuals from the latent slow (*L*_*s*_) slow compartment can lead to asymptomatic TB (*S*_*c*_). Progression from asymptomatic TB (*S*_*c*_) to symptomatic TB (*A*) occurs over time. Both asymptomatic and symptomatic TB disease affected individuals were assumed to be equally infectious, consistent with evidence suggesting that asymptomatic cases, despite lacking classical symptoms, play an important role in sustaining TB spread (9).

Once symptomatic, individuals may either seek care or not seek care (i.e. remain undiagnosed) in which case they are sent to the *O* compartment. Individuals seeking care may present to either public *G* or private *P* providers for diagnosis. Only a certain proportion of individuals seeking care are diagnosed with TB who are sent to the compartment *D*, with misdiagnosed individuals returning to the O compartment. The diagnosed individuals upon treatment initiation enter *T*_*I*_. A proportion of individuals who received a correct diagnosis may fail to initiate treatment and are assumed to transition to the O compartment. Among those who start treatment, outcomes include treatment success *T*_*S*_, loss to follow-up *F*, or death. Individuals in *O* compartment may re-enter the care pathway by seeking or re-seeking care at any point of time.

Successfully treated individuals were assumed to have a low relapse risk and entered the *T*_*S*_ compartment, whereas those who did not complete treatment were assumed to have a higher relapse risk and entered the *F* compartment. A proportion of individuals from these groups can have stabilised risk of relapse after a period of time and enter the *R* compartment. In addition, all the three groups at risk of relapse were assumed to be at risk of reinfection, upon which they re-entered the latent fast compartment *L*_*f*_.

The complete framework consists of 13 compartments, capturing the pathway from infection acquisition and disease progression to treatment completion, reinfection, and relapse (Figure 1). All compartments are expressed as proportions of the population. The model does not distinguish between adult and paediatric TB, pulmonary and extrapulmonary TB, or account for co-infections and drug resistance. Demographic balance is maintained by equating birth into the uninfected compartment with deaths due to natural and TB-related causes across all compartments.

**Figure 1:**
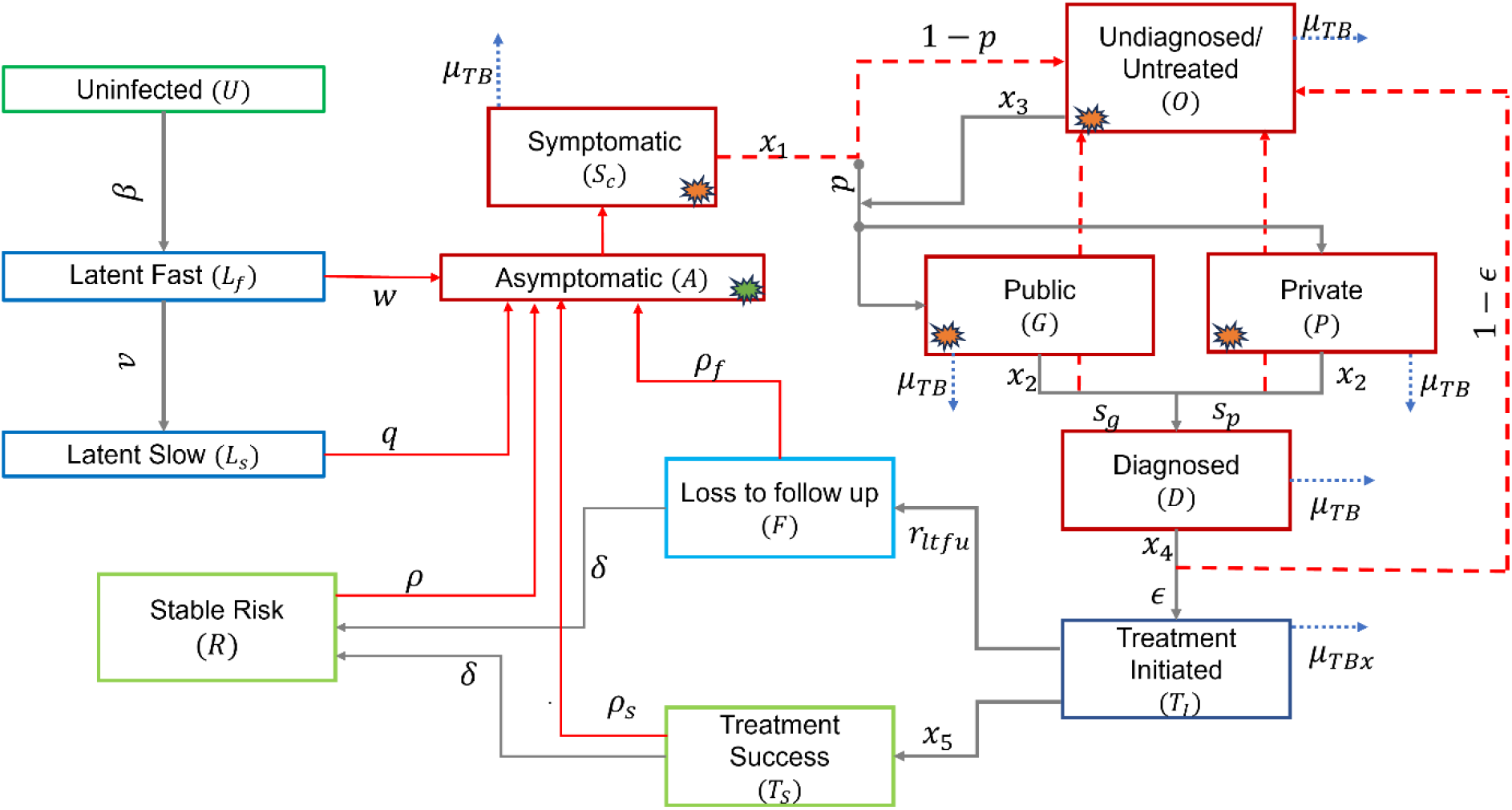
Schematic representation of the model. For simplicity, certain rates such as self-cure, background mortality, and exogenous reinfection are not displayed. Red compartments contribute to the force of infection, green compartments represent individuals who are susceptible or recovered, and blue compartments capture the intermediate stages of infection and recovery. Red solid lines indicate the pathway to disease, the red dashed lines denote attrition during care-seeking, diagnosis, treatment initiation. Targets for symptom-based interventions are compartments with orange stars and targets for symptom-agnostic interventions are orange and green stars

### Model Calibration

The model was calibrated to the data listed in (Table 1) using an adaptive Bayesian Markov chain Monte Carlo (MCMC) approach (10). We used the TB notification rate from 2019 (11), representing the most recent pre-pandemic data, and from 2023 (12), reflecting the stabilization of programmatic systems following the disruptions caused by the COVID-19 pandemic. Estimates of TB prevalence, the proportion of prevalent TB that is asymptomatic, and the prevalence-to-notification (P:N) ratio were obtained from the National TB Prevalence Survey (2). Mortality data was sourced from the Global TB Report (1). The complete data table with their values is listed in Table 1. Posterior densities were constructed for the calibration targets, with model proportions represented by beta distributions and rates by log-normal distributions. Parameters were adjusted to match the observed uncertainty intervals. We assumed sufficiently wide priors to allow effective exploration of the parameter space. Posterior densities were taken to be proportional to the product of likelihoods and priors under the Bayesian framework. We calculated the log-posterior density, therefore taking a sum of the log-probability distributions of each of the individual likelihood components.

**Table 1:**
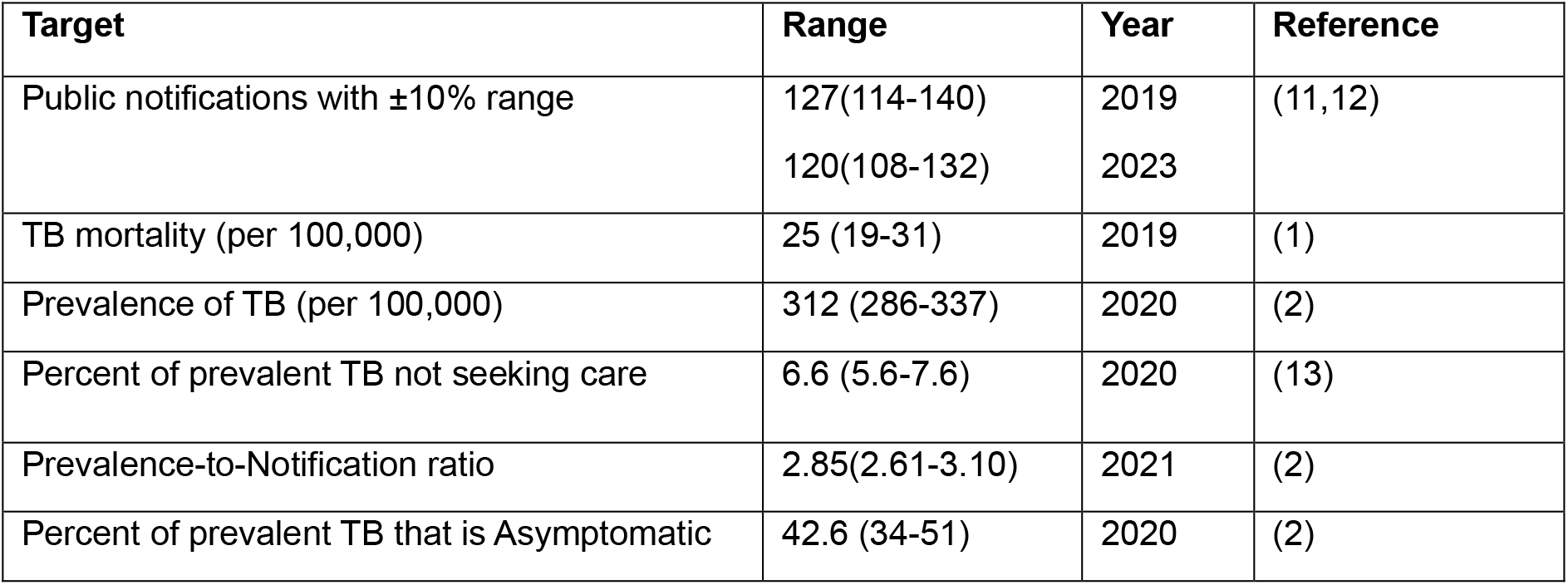
Summary of calibration targets used in the model. Where uncertainty bounds were not directly reported in the source data, symmetric uncertainty was applied depending on the data type.

| Target | Range | Year | Reference |
| --- | --- | --- | --- |
| Public notifications with $\pm 10\%$ range | 127(114-140) | 2019 | (11,12) |
|  | 120(108-132) | 2023 |  |
| TB mortality (per 100,000) | 25 (19-31) | 2019 | (1) |
| Prevalence of TB (per 100,000) | 312 (286-337) | 2020 | (2) |
| Percent of prevalent TB not seeking care | 6.6 (5.6-7.6) | 2020 | (13) |
| Prevalence-to-Notification ratio | 2.85(2.61-3.10) | 2021 | (2) |
| Percent of prevalent TB that is Asymptomatic | 42.6 (34-51) | 2020 | (2) |

Sampling was initiated by generating 5,000 parameter sets using Latin Hypercube Sampling, from which the three sets with the highest posterior densities were selected as starting points for independent MCMC chains. Half of the iterations were discarded as burn-in, and 250 samples were drawn from the remaining iterations. Model outputs were summarised using the 50th percentile as the central estimate and the 2.5th and 97.5th percentiles to define the 95% Bayesian credible intervals. Trace plots of the MCMC sampling are shown in (Figure 1 (A) and (B), supplementary), calibration results are presented in (Figure 1 (C) and (D) supplementary).

### Intervention scenarios

We evaluated multiple intervention scenarios by framing them under two complementary perspectives: targeted approach and generalised approach. The targeted approach serves as an umbrella for efforts that can be implemented at the system level, such as accelerated active case finding, nutritional support, and health system strengthening. Health system improvements were incorporated by progressively increasing the probability of correct diagnosis in both the public and private sectors denoted by *s*_*g*_ and *s*_*p*_, respectively, from their baseline estimates of 0.56(0.19-0.88) and 0.26(0.10-0.65) respectively to 0.9 by 2030, alongside a 10% annual increment in the estimated rate of diagnosis *x*_2_. Nutritional intervention was implemented by scaling up the rate of stabilisation v annually, and was calibrated to reproduce a population-level incidence decline comparable to that observed in the RATIONS trial (14). Active case finding (ACF) as an intervention was modelled by transferring individuals from the undiagnosed compartments directly to the treatment initiation compartment, thereby substantially reducing the time to diagnosis.

In contrast the generalised approach explored the interventions in terms of the transmission rate *β*. Transmission rate summarises the combined impact of improvements in the social, behavioural and economic factors together with TB control programs in preventing new infections. These efforts can be modelled as an annual decline in *β*(15,16). In both the approaches the interventions were assumed to be in place till the year 2030 starting from 2025. The baseline scenario for all the intervention approaches is the no intervention case.

## RESULTS

The model was simulated to estimate TB incidence and mortality rates per 100,000 population between 2000 and 2025. For 2024, the estimated incidence was 180 (164-197) and the mortality rate was 24 (19–30) per 100,000 population (Figure 2) broadly consistent with WHO estimates of incidence and mortality of 187(160-218) and 21(16–26) per 100,000. Posterior distributions of the parameters estimated during calibration are shown in (Figure 2, supplementary), with central value and 95% credible intervals listed in (Table 1, supplementary). A sensitivity analysis using partial rank correlation coefficients of the input parameters was performed with respect to baseline incidence and mortality of 2024 and the time period of intervention scenarios (figure 4, supplementary).

**Figure 2:**
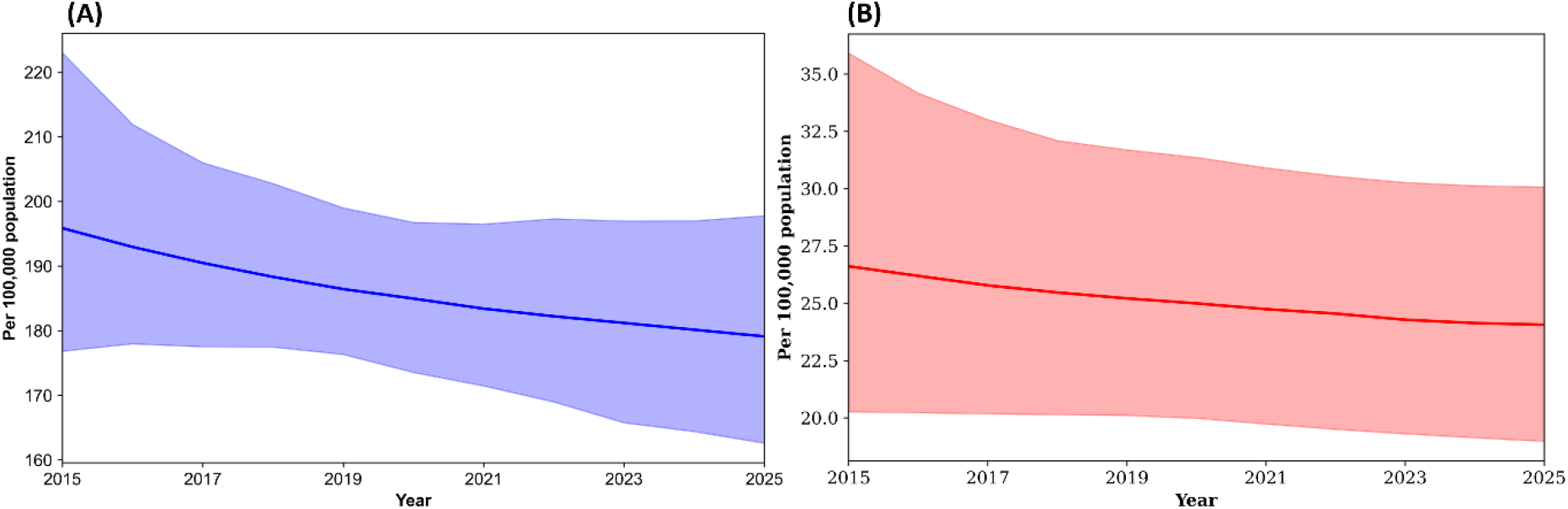
Baseline model estimates in the absence of any interventions discussed in the manuscript. The estimated incidence rate between 2015 and 2025 is shown in panel (A) and the mortality rate in panel (B). The thick line represents the central estimate, while the shaded region indicates the 95% uncertainty interval.

### Force of Infection

The force of infection, representing the annual number of new infections to which susceptible individuals are exposed, depends both on transmission parameters and on the relative sizes of infectious compartments. Comparison of these compartments revealed that asymptomatic individuals were the largest contributors 51(40-61) % to transmission (Figure 3). This finding underscores the critical role of identifying and treating asymptomatic TB.

**Figure 3:**
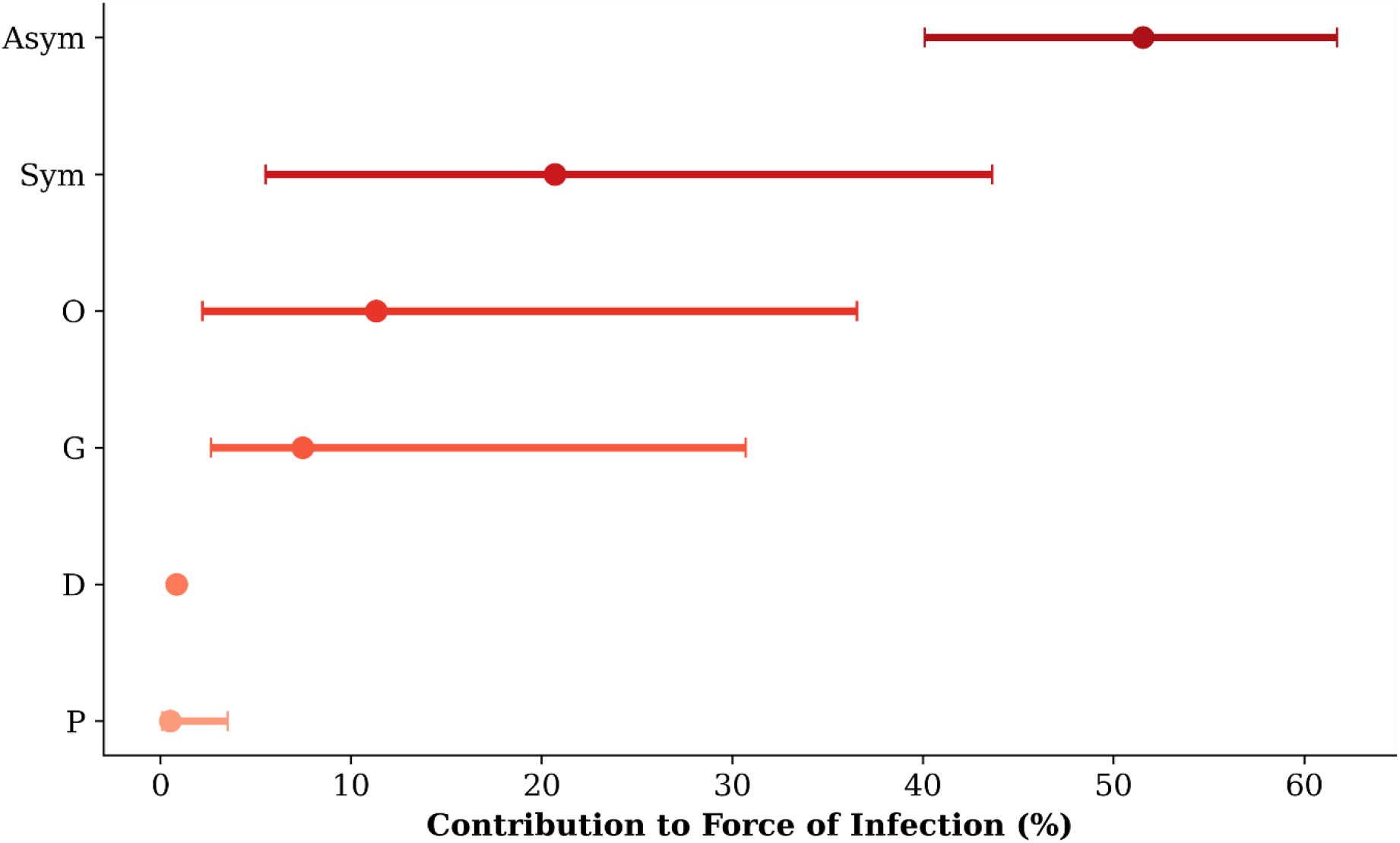
The x-axis represents percentage contribution. The compartments are labelled as Asym = asymptomatic, Sym = symptomatic, O = not seeking care, G = public health facility, P = private health facility, and D = diagnosed.

### Nutritional support

Undernutrition is a major risk factor for both the development of active TB and poor treatment outcomes (17–19). Interventions targeting this determinant are therefore critical for TB control. The RATIONS trial (14) demonstrated that providing nutritional support to household contacts of patients with TB reduced TB incidence by 40% over a period of two years. To translate these cohort-level findings into a population-level model, we represented the effect of nutritional support as an improvement in the stabilisation rate v from the latent fast to the latent slow state. For the full population model, achieving a reduction of TB incidence by 40% implied an increase in the stabilisation rate by 42% relative to the baseline (Figure 3, supplementary). However, taking into account that the RATIONS trial focused specifically on household contacts of patients with TB, we adopted a conservative approach and considered two scenarios of an increased stabilisation rate by 10% and 25% compared to the baseline model.

Under a 10% annual improvement in rate of stabilisation, TB incidence declined by 28(25–31) % to 128(118-138) cases and mortality by 20(16–25) % to 20(16–25) deaths by 2030. A more ambitious 25% annual improvement scenario resulted in a 51 (47-56) % reduction in incidence to 87(83-91) cases and a 34(32-38) % reduction in mortality to 16(13–19) deaths (Figure 4 (A) and (B)).

**Figure 4:**
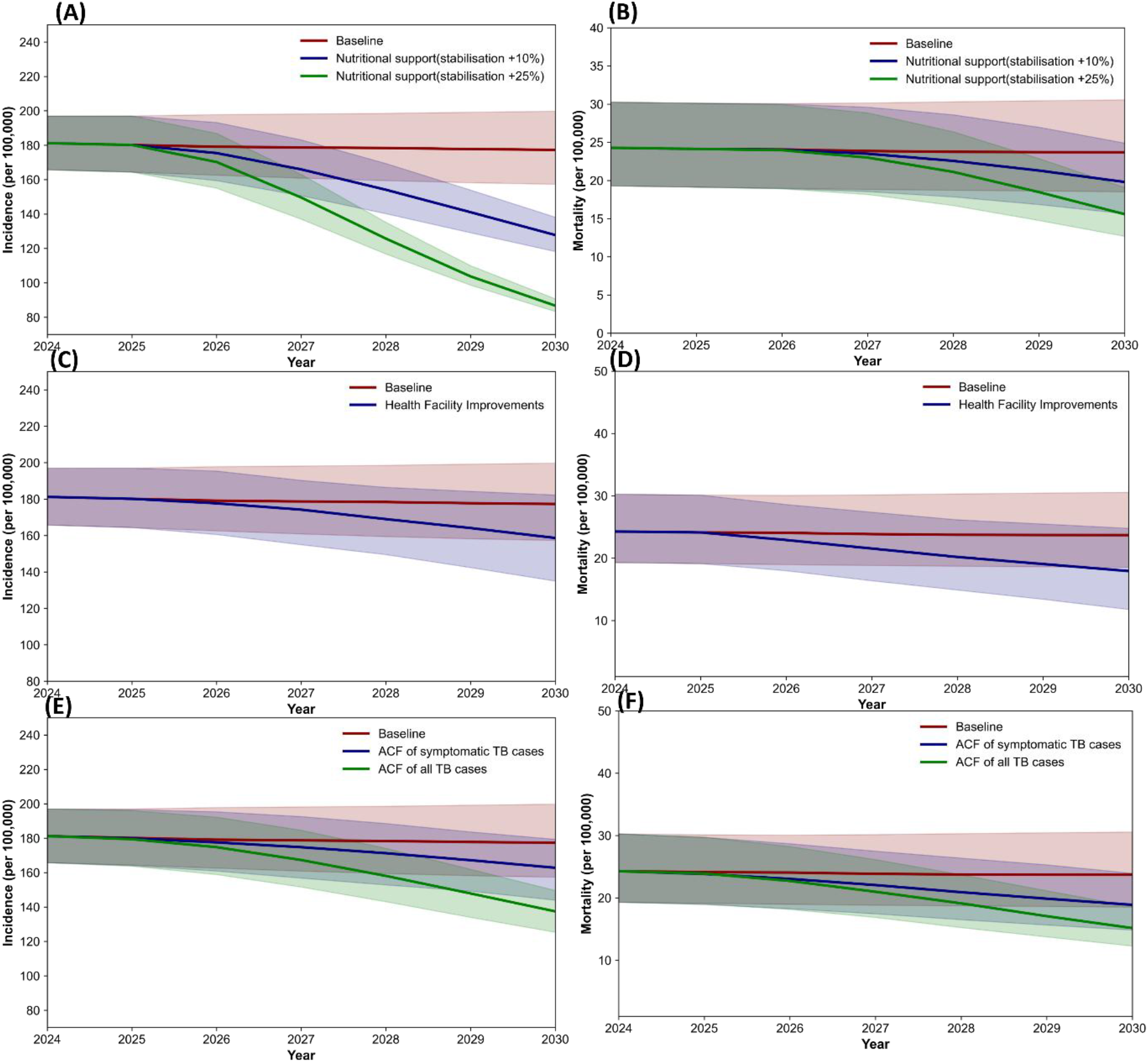
Each row represents a specific intervention with incidence (left) and mortality rate per 100,000 population(right). Row 1: Nutritional support, Row 2: Health facility improvements, Row 3: Active case finding.

### Health system strengthening

Strengthening the health systems, particularly the efficiency of private and public sector diagnosis, is critical to reducing undiagnosed TB. This scenario led to a 11(9–14) % % reduction in incidence rate with incidence reaching 159(135-182) per 100,000 while mortality rate declined by 24(18-36) % with deaths per 100,000 declining to 18(12–25) in the year 2030 (Figure 4 (C) and (D)).

### Accelerated active case finding

Two active case-finding (ACF) strategies were evaluated: symptom-based and symptom-agnostic. Symptom-agnostic approaches, such as chest X-ray with computer-aided detection, can identify asymptomatic TB cases that are missed by symptom-based screening (2). An ACF intervention was incorporated into the model, assuming it would be implemented in addition to existing programmatic activities.

The intensity of ACF was assumed to improve annually such that, by 2030, 40% of all TB cases would be identified by this approach. In the model, the detection rate of undiagnosed TB cases was gradually scaled up from 2025 to achieve 40% detection of both symptomatic and asymptomatic cases by 2030.

Under this scale-up scenario, symptom-based ACF led to an 8(9–10) % reduction in incidence and a 20(19–21) % reduction in mortality. The estimated incidence and mortality rates after the implementation of this scenario are 163(144-179) and 19(15–24) per 100,000. Symptom-agnostic ACF achieved larger declines of 22(20–25) % in incidence and 36(34-39) % in mortality with incidence and mortality reaching 137(125-150) and 15(12–19) per 100,000 respectively by 2030 (Figure 4 (E) and (F)).

### Multi-sectoral strategy

To achieve India’s TB elimination goals of reducing Incidence by 80% and mortality by 90% compared to 2015, coordinated multi-sectoral action is essential. Simulating a combined intervention across all three domains discussed above produced a 60 (58-62) % decline in incidence and a 66(65-67) % decline in mortality leading to 71(66-75) cases and 8(6–10) deaths per 100,000 in 2030 (Figure 5 (A) and (B)). These findings highlight that isolated interventions are unlikely to suffice; instead, sustained-integrated efforts across health, nutrition, and social systems are critical to translate modelled gains into real-world progress toward TB elimination.

**Figure 5:**
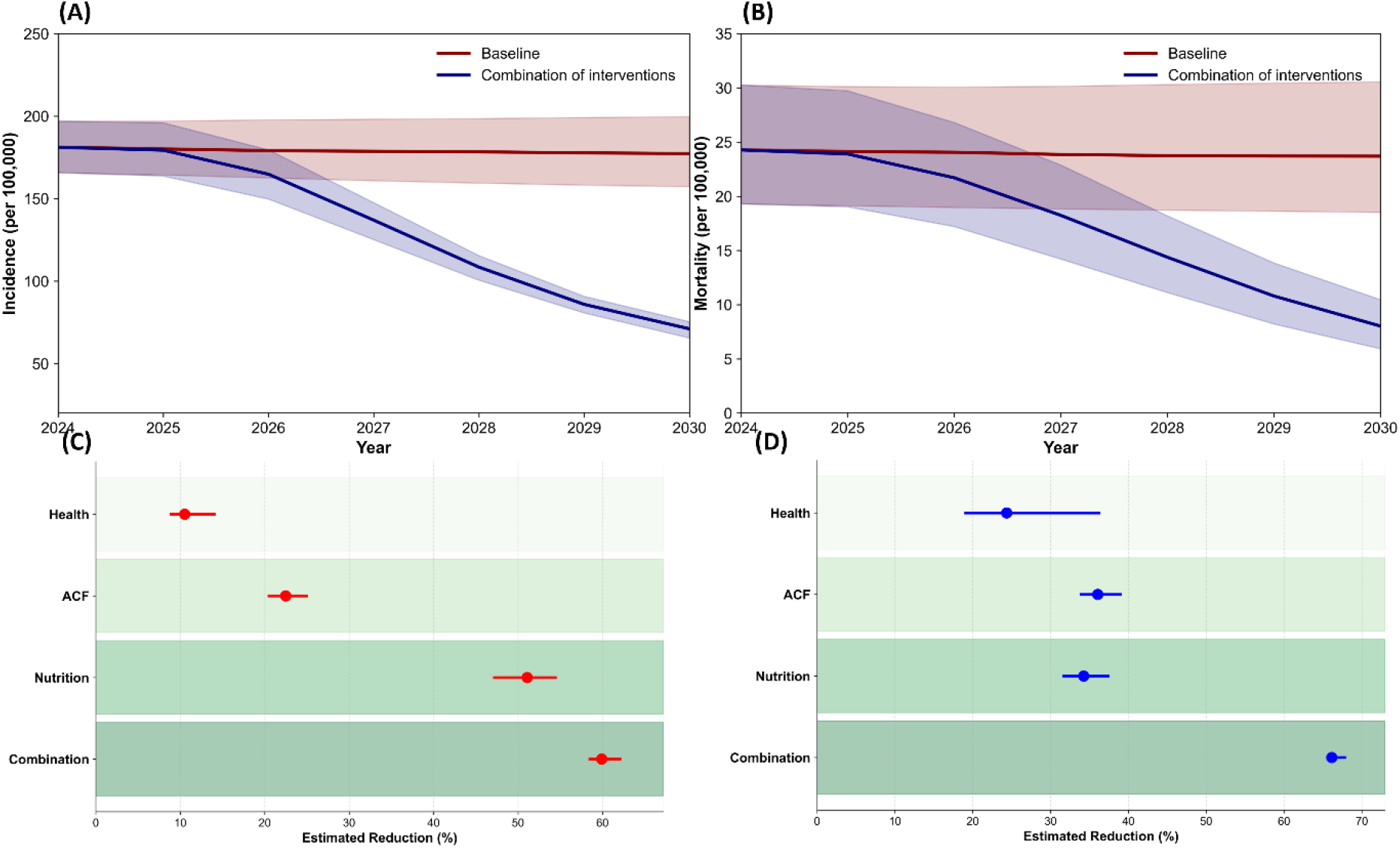
Top row: Impact of multi-sectoral interventions. Bottom row: Comparison of decline in incidence and mortality when different intervention strategies are implemented Nutrition refers to the nutritional support provided to TB infected persons, resulting in 25 percent increase in the stabilisation rate; ACF refers to active case finding of all the TB cases; Health refers to improvements in the health facilities. Combination refers to a combination of all three interventions.; For a detailed description of each strategy the reader is advised to refer to the text.

### Generalised approach

Transmission rate describes how quickly an infection spreads from infectious individuals to susceptible individuals in a population. An annual 10% reduction in transmission rate, as discussed in the methods section, led to a 29(26–32) % decline in incidence leading to 126(117-136) per 100,000 population in 2030, and a 17(16–20) % decline in mortality to 20(15–25) per 100,000 over the same period (Figure 6 (A) and (B)).

**Figure 6:**
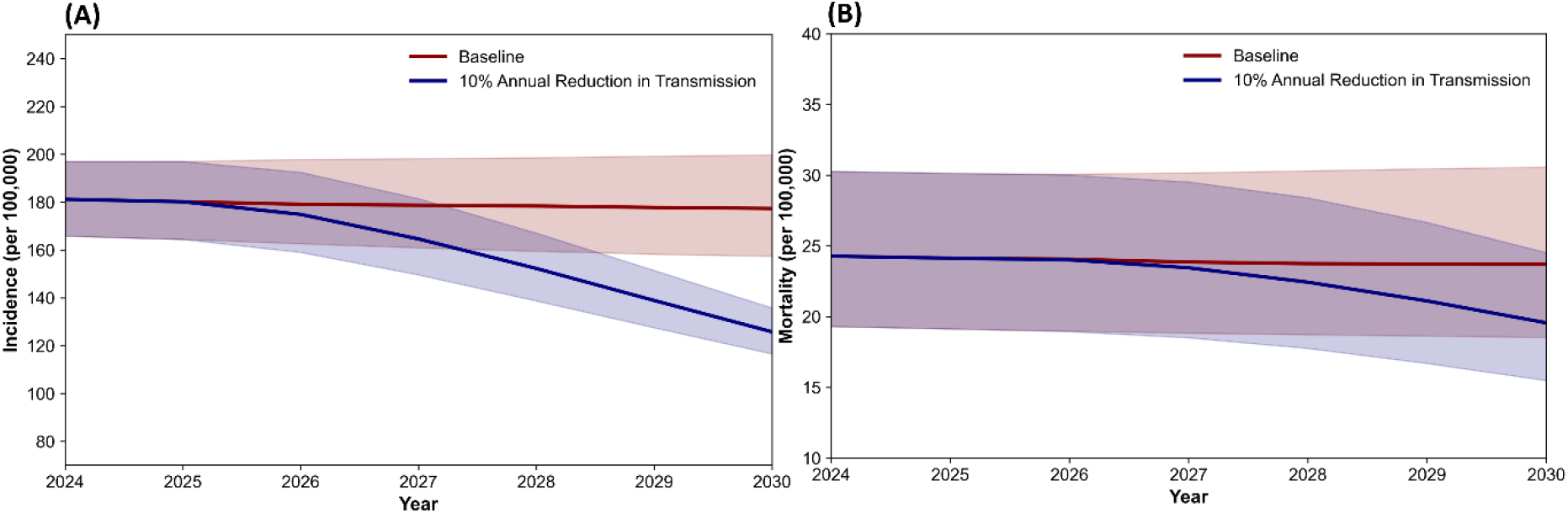
The impact of reduction in transmission on incidence and mortality

## DISCUSSION

In this modelling study, we developed a TB transmission model that yielded two critical insights for informing programmatic policy. First, our model provides quantitative estimate that asymptomatic-infectious (aTB) individuals are responsible for over half of all new infections. Second, our projections demonstrate that while individual interventions make progress, substantial epidemiological gains are possible only with a comprehensive, multi-component strategy that is truly multi-sectoral.

The significance of this large aTB group is increasingly recognized and holds programmatic importance. Our estimate that individuals with asymptomatic disease contribute 51(40-61) % of new infections in India is consistent with emerging evidence. This finding is biologically plausible, as studies have shown that the exhalation of infectious aerosols is not necessarily dependent on the presence of symptoms like coughing (20).

Our model’s primary contribution is its structure, which bridges this new understanding of pathogenesis and natural history with a strategic focus on programmatic as well as health system action. Its unique value lies in the development of an asymptomatic compartment with a detailed, multi-pathway care cascade that explicitly includes India’s large public and private sectors, as well as the substantial proportion of individuals who do not seek care at all, a structure calibrated directly to the recent national prevalence survey data. This allows for a realistic evaluation of how interventions interact with real world health-seeking behaviours.

The model also shows that a structured shift from passive to active case finding is essential especially in the light of asymptomatic individuals contributing to more than 50% of all transmission. This is also validated by our projections which estimate that symptom-agnostic screening has a far greater potential to interrupt community transmission than traditional symptom-based approaches. This is not a new concept: historical campaigns in now low-burden countries successfully used mass chest X-ray screening to achieve population-level impacts (21). Modern tools like portable X-ray with AI-assisted interpretation make this approach more feasible today, which India has already begun leveraging in its recent national campaign (22).

Our model was specifically built to evaluate the impact of intervention scenarios that act upon the complex natural history of TB and care cascade. This focus on targeted strategy is complementary to other major modelling efforts in India such as addressing under-nutrition as a preventive measure (19), on introducing TB vaccines both newer vaccines and BCG revaccination (23), private sector engagement and health system strengthening efforts (13,24) as well as strengthening case finding efforts through active case-finding (25). However, as with all modelling studies, a key limitation is that the projected impact reflects efficacy under idealised assumptions, and the real-world effectiveness of these interventions may be lower due to implementation challenges.

The success of any screening strategy is contingent upon strengthening the entire cascade of care, which remains a challenge in the country (26). For instance, an idealised assumption of achieving a 90% diagnostic accuracy in the private sector which is fragmented and unregulated is highly ambitious but achievable through strengthening the health system and care cascade components within the private sector. Similarly, modelling interventions as a reduction in the transmission parameter β is a simplified effective picture as it does not explicitly capture the effects of individual interventions, behavioural changes, or structural determinants that influence transmission dynamics, neither does it address the deep-seated TB-related stigma (27) and corresponding socio-economic determinants (28) that sustain transmission.

Despite these simplifications, the model offers a more comprehensive and realistic assessment of intervention impact than methods limited to a single domain. In doing so, it offers a stronger foundation for designing strategies that reflect the diverse drivers of TB in India. A major limitation of the work is the lack of age-structure (29,30) a key factor in TB epidemiology. It also doesn’t distinguish between key risk groups or different forms of TB. An important question which we aim to address in the future work is the incorporation of age-stratification which would provide key insights into the impact of asymptomatic infectious individuals on the transmission dynamics and shape India’s efforts towards ending TB in the country.

To conclude, our study provides compelling evidence that the burden of aTB is high in the country. To translate modelled projections into epidemiological gains, a dynamic, multi-sectoral, and integrated approach for ending TB is a necessity. This requires strengthening health systems, making nutritional support a standard of care, and addressing the social determinants of health and multi-sectoral accountability for achieving the population-level impact by 2030.

## Data Availability

All data produced in the present work are contained in the manuscript

## Acknowledgement

The authors thank Dr. Rajiv Bahl, Director General, Indian Council of Medical Research (DG-ICMR) for his valuable insights, comments and guidance.

## DECLARATIONS

### Ethics approval and consent to participate

Not applicable.

### Availability of data and materials

All data used in the model is publicly available with their sources mentioned wherever required. The software and package utilised are open access and available online.

### Author contributions

YR, MM, SM contributed in the conceptualisation and design of the work. YR conducted data collection and formal analysis of the work. YR wrote the first draft of the manuscript with assistance from RP. The final draft of the manuscript was revised and approved by MM and SM.

### Disclaimer

The work/opinion is based on research findings by the authors and not the opinion of the government.

