## Supplementary Figures and Tables for "Assessing the Impact of Interventions on Tuberculosis Control: India Based Modelling Framework"

Table of Contents

Supplementary Material……………………………………………………………………1

Text S1. Details of Model and sensitivity analysis

Model Equations………...……………………………………………………..2

Sensitivity Analysis……..………………………………………………………8

Supplementary Figures

Supplementary Figure 1………………………………………….……………….4

Supplementary Figure 2…………………………………………………………..6

Supplementary Figure 3…………………………………………………………..8

Supplementary Figure 4…………………………………………………………..9

Supplementary Table

Supplementary Table 1…………………………..……………………………….5

*Model Equations*

The compartment $U$ represents the uninfected population. Individuals enter this compartment through birth and exit at a rate determined by the force of infection $\lambda$ and natural mortality $\mu$

$$\frac{dU}{dt} = b - \lambda U - \mu U$$

Upon infection, individuals enter the latent fast compartment $L_f$ either through primary infection or reinfection. From $L_f$ individuals may progress rapidly to asymptomatic TB $S_c$ or stabilize into the latent slow compartment $L_s$. A background mortality rate μ representing mortality due to natural causes is included in the compartments $U, L_f, L_s$.

$$\frac{dL_f}{dt} = \lambda U + \lambda(1 - h)(L_s + T_s + F + R) - (v + w)L_f - \mu L_f$$

$$\frac{dL_s}{dt} = vL_f - qL_s - \lambda(1 - h)L_s - \mu L_s$$

Individuals progress from asymptomatic to symptomatic TB $A$ at the rate $\alpha$, with untreated TB mortality $\mu_{tb}$ included in $A$ .

$$\frac{dS_c}{dt} = wL_f + qL_s + \rho_s T_s + \rho_f F + \rho_r R - (\alpha + \sigma)S_c - \mu S_c$$

$$\frac{dA}{dt} = \alpha S_c - (x_1 + \sigma)A - \mu_{tb} A$$

Incidence was defined as the number of new active TB cases in a given year entering the asymptomatic compartment. Subclinical TB represents an early stage of the disease with no classical signs and symptoms associated with the clinical stage of the disease. TB-related mortality is generally associated with more advanced stages of the disease; therefore, only natural mortality is applied to the asymptomatic compartment. A proportion $p$ of symptomatic individuals seek care through public $G$ or private $P$ healthcare providers. The parameter $p_g(t)$ represents the scale-up of public healthcare and captures its impact on changes in TB notification rates. We allowed this parameter to change in a linear way between 2000 and 2010 capturing the scale up of DOTS services in this period leading to people choosing public sector on each care seeking visit, thereby impacting the public sector notifications over time. The parameter $x_1$ denotes the rate at which individuals seek initial care after symptom onset. Individuals in the $O$ compartment may re-enter the care pathway by seeking or re-seeking care at a later stage with a rate $x_3$ . The diagnosis rate is given by $x_2$.

$$\frac{dO}{dt} = x_1(1 - p)A + x_2\left(1 - s_g\right)G + x_2\left(1 - s_p\right)P + x_4(1 - \epsilon)D - (x_3 + \sigma)O - \mu_{tb}O$$

$$\frac{dG}{dt} = p_g(px_1A + x_3O) - (x_2 + \sigma)G - \mu_{tb}G$$

$$\frac{dP}{dt} = \left(1 - p_g\right)(px_1A + x_3O) - (x_2 + \sigma)P - \mu_{tb}P$$

The probability of a successful diagnosis at the public and private healthcare provider is given by $s_g$ and $s_p$ respectively. Diagnosed individuals enter compartment $D$, a fraction $\epsilon$ initiate treatment at the rate $x_4$ moving into the compartment $T_I$ .

$$\frac{dD}{dt} = s_gx_2G + s_px_2P - (x_4 + \sigma + \mu_{tb})D$$

$$\frac{dT_I}{dt} = \epsilon x_4D - x_5T - r_{ltfu}T - \mu_{tbx}T$$

TB-related mortality was calculated as the sum of deaths among untreated cases and deaths occurring during TB treatment. Treatment completion occurs at the rate $x_5$ and treatment is interrupted at the rate $r_{ltfu}$. Treatment completion moves the patients to the $T_s$ compartments with a lower risk of relapse while interruptions move the patients into the $F$ compartment with a higher risk of relapse. Long term recovery and stabilized risk is modelled through the $R$ compartment.

$$\frac{dT_s}{dt} = x_5T - \left(\lambda(1 - h) + (\rho_s + \delta)\right)T_s - \mu T_s$$

$$\frac{dF}{dt} = r_{ltfu}T + \sigma(S_c + A + O + P + G + D) - \big(\lambda(1 - h) + \rho_f + \delta\big)F - \mu F$$

$$\frac{dR}{dt} = \delta(T_s + F) - (\lambda(1 - h) + \rho_r + \mu)R$$

The force of infection $\lambda$ is given by

$$\lambda = \beta(S_c + A + O + G + P + D)$$

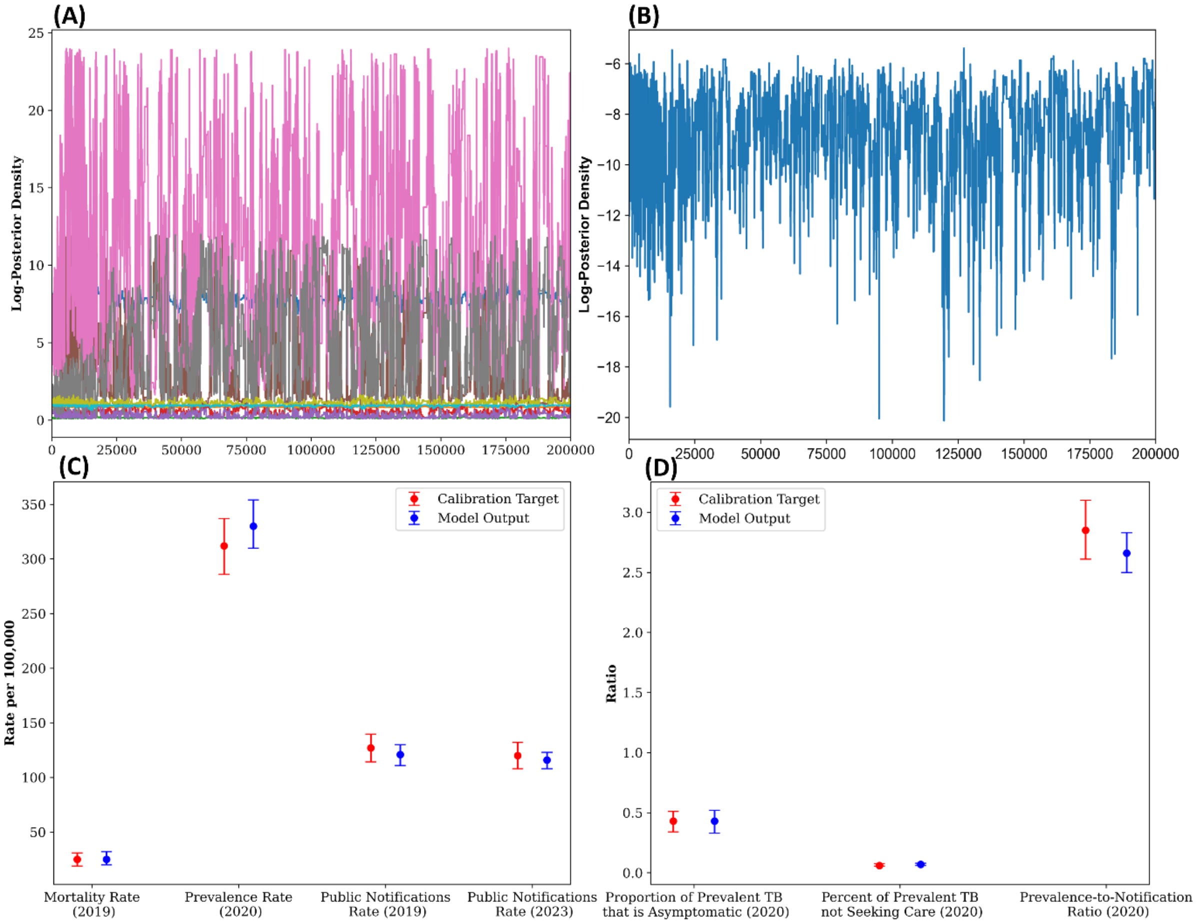


*Figure 1: Top row: MCMC trace plots Bottom row: Comparison of model fit with data, Numbers in brackets refer to the year of the data used for calibration.*

| Symbol | Meaning | Value | Source |
|---|---|---|---|
| $\beta$ | Annual infections per TB case | 7.8 (7.2-8.5) | Estimated; prior U[0,30] |
| $v$ | Per-capita annual rate of stabilisation from $L_f$ to $L_s$ | 0.872 | (31) |
| $w$ | Per-capita annual of fast progression from $L_f$ to $S_c$ | 0.0826 | (31) |
| $q$ | Per-capita annual rate of slow progression from $L_s$ to $S_c$ | 0.0006 | (31) |

| | | | |
|---|---|---|---|
| $\alpha$ | Per-capita annual rate of developing symptoms | 1.1(0.91-1.5) | Estimated; prior U[0,12] |
| $\sigma$ | Per-capita annual rate of self-cure | 1/6 | (32) |
| $x_1$ | Rate of initial care seeking ($yr^{-1}$) | 2.5 (1.0-10) | Estimated; prior U[0.1,12] |
| $x_2$ | Rate of diagnosis ($yr^{-1}$) | 11.4 (2.4-23.44) | Estimated; prior U[1,24] |
| $x_3$ | Rate of entering/re-entering care pathway ($yr^{-1}$) | 5.4 (0.87-11.2) | Estimated; prior U [0.1,12] |
| $x_4$ | Rate of treatment initiation ($yr^{-1}$) | 52 | Assumed (1 week) |
| $x_5$ | Rate of treatment completion ($yr^{-1}$) | 2 | 6-month treatment duration |
| $p_g$ | Proportion seek Ing care in public | 0.94(0.80–0.98) | Estimated; prior U[0,0.98] |
| $p$ | Proportion seeking care | 0.84(0.81–0.88) | Estimated; prior U [0.1,0.9] |
| $s_g$ | Probability of correct diagnosis in public sector | 0.56(0.18-0.88) | Estimated; prior U [0.1,0.9] |
| $s_p$ | Probability of correct diagnosis in private sector | 0.26(10-0.65) | Estimated; prior U [0.1,0.9] |
| $\epsilon$ | Proportion initiated on treatment | 0.95 | Assumed |
| $r_{ltfu}$ | Rate of treatment interruptions ($yr^{-1}$) | 0.133 | 6% post-treatment LTFU |
| $h$ | Protection from reinfection ($yr^{-1}$) | 0.5 | (33) |
| $\rho_s$ | Per-capita annual relapse rate after successful treatment | 0.032 | (34) |
| $\rho_f$ | Per-capita annual relapse rate after treatment interruptions | 0.14 | (34) |
| $\rho$ | Per-capita annual relapse rate after two years | 0.0015 | (34) |
| $\delta$ | Rate of stabilization to long term risk ($yr^{-1}$) | 0.5 | (35) |
| $\mu$ | Per-capita annual natural mortality rate | 1/72 | Fixed |
| $\mu_{tb}$ | Per-capita annual TB mortality rate (untreated) | 0.15 (0.11–0.23) | Estimated; untreated CFR 29(20–40) % |
| $\mu_{tbx}$ | Per-capita annual TB mortality rate during treatment | 0.0888 | CFR 4% during treatment |

70 *Table 1: Posterior ranges of the estimated parameters*

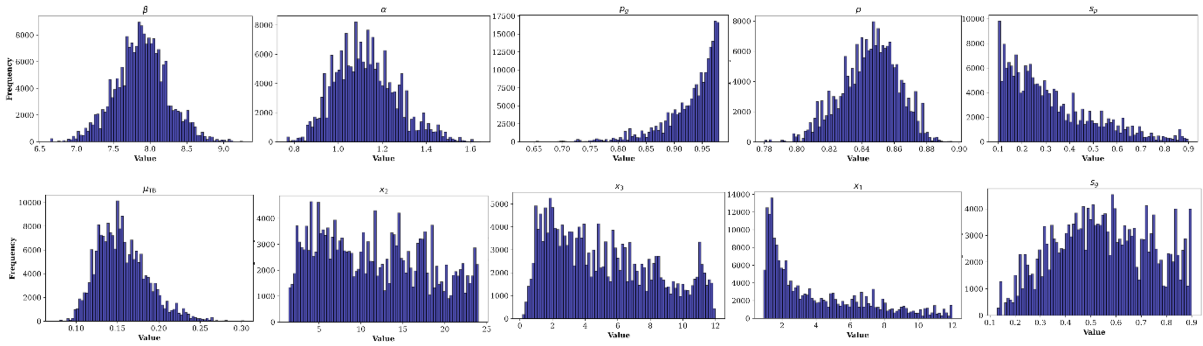

Figure 2: Posterior distribution of the estimated parameters

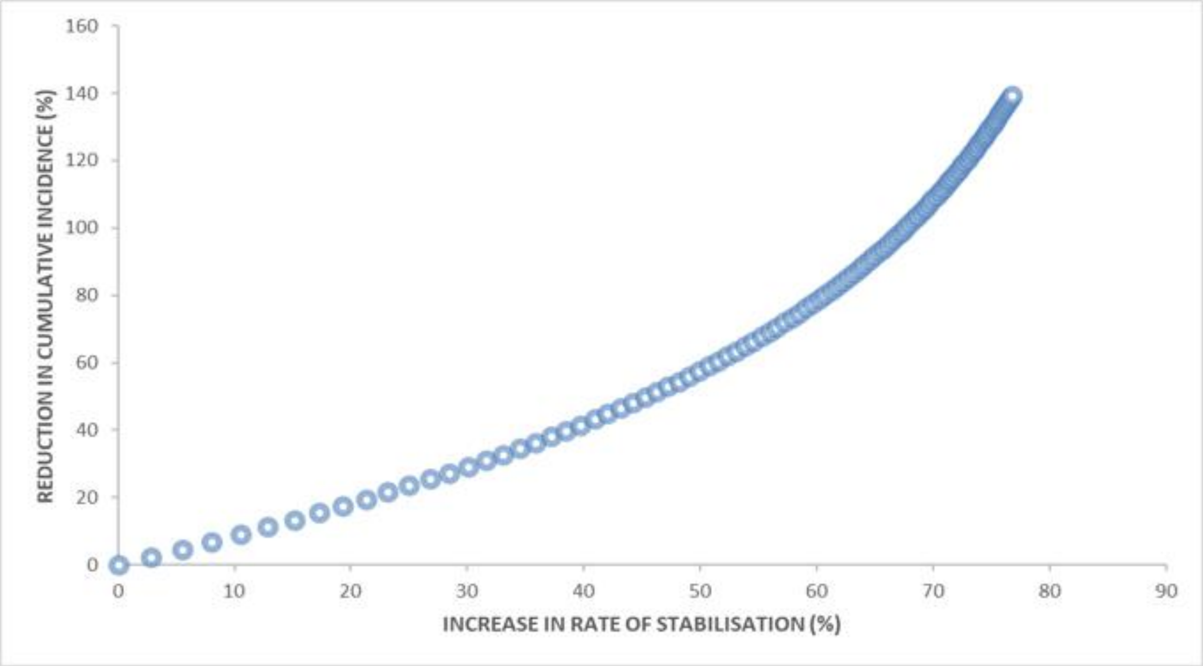


Figure 3: The impact of annual increment in rate of stabilisation resulting in incidence reductions

### Sensitivity Analysis

To identify the effect of the estimated parameters on the modelled baseline incidence and mortality for the year 2024, we performed a sensitivity analysis using the partial rank correlation coefficients (PRCC) (figure 4 (a) and (b), supplementary). To examine how the parameter influence evolved over time, we computed PRCC for two time points: (i) at the start of the intervention period 2025 and (ii) at the end of the intervention period 2030. Changes in the PRCC magnitude and direction helped to identify the parameters whose relative importance shifted over the course of intervention. Parameters with larger PRCC are interpreted as having stronger influence on the outcomes of incidence and mortality figure 4 (c) and (d), supplementary).

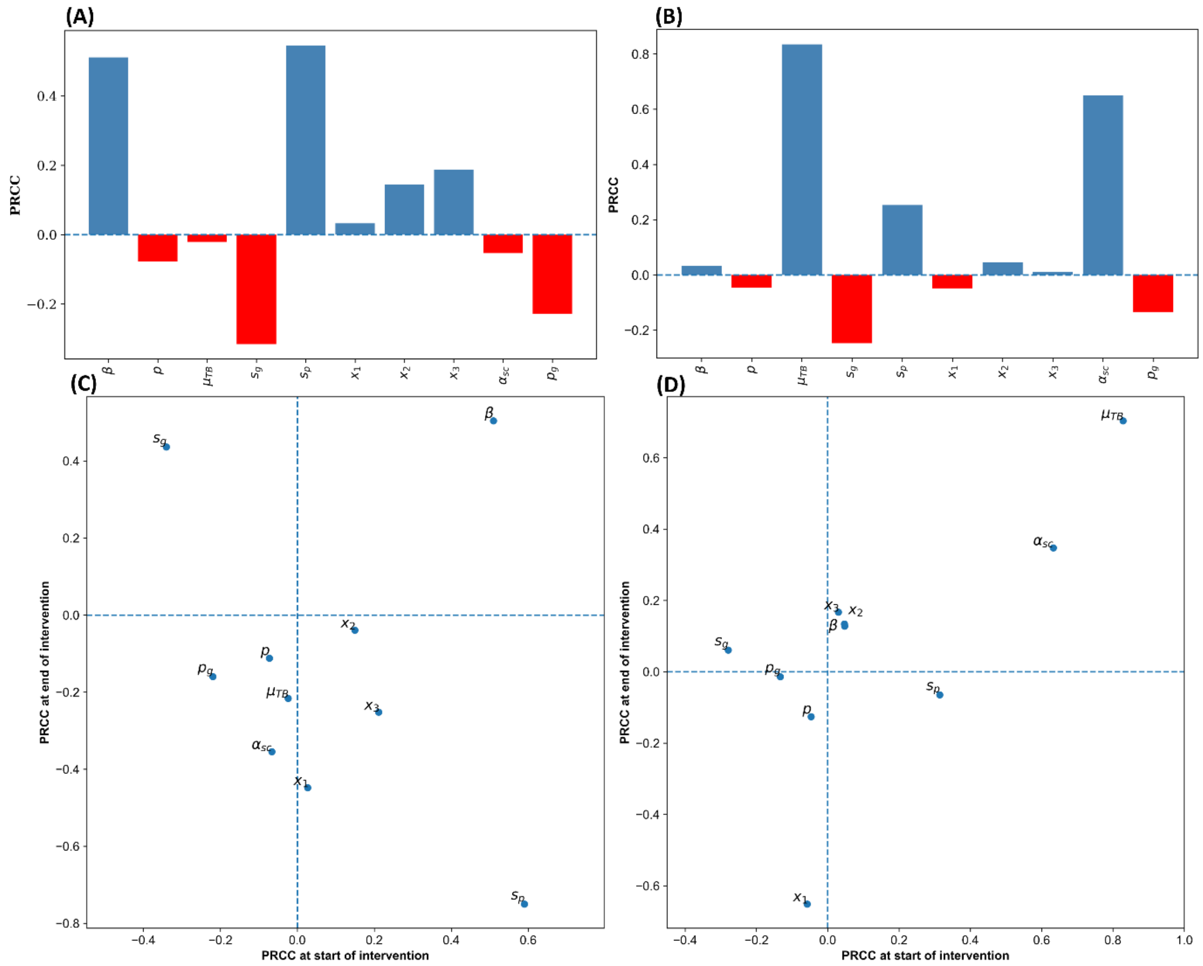


*Figure 4: Sensitivity of modelled incidence and mortality to parameter uncertainty. Panels (A) and (B) show the PRCC values for incidence and mortality with respect to model parameters. Panels (C) and (D) compare PRCC values at the start and end of the intervention period for incidence and mortality, respectively, illustrating how parameter importance changes over time. Parameters with PRCC values farther from zero have a stronger influence on the outcomes. Horizontal and vertical dashed lines indicate zero correlation.*
